# SARS-CoV-2 viral load peaks prior to symptom onset: a systematic review and individual-pooled analysis of coronavirus viral load from 66 studies

**DOI:** 10.1101/2020.09.28.20202028

**Authors:** Amy E. Benefield, Laura A. Skrip, Andrea Clement, Rachel A. Althouse, Stewart Chang, Benjamin M. Althouse

## Abstract

**Background:** Since the emergence of COVID-19, tens of millions of people have been infected, and the global death toll approached 1 million by September 2020. Understanding the transmission dynamics of emerging pathogens, such as SARS-CoV-2 and other novel human coronaviruses is imperative in designing effective control measures. Viral load contributes to the transmission potential of the virus, but findings around the temporal viral load dynamics, particularly the peak of transmission potential, remain inconsistent across studies due to limited sample sizes.

**Methods:** We searched PubMed through June 8th 2020 and collated unique individual-patient data (IPD) from papers reporting temporal viral load and shedding data from coronaviruses in adherence with the PRISMA-IPD guidelines. We analyzed viral load trajectories using a series of generalized additive models and analyzed the duration of viral shedding by fitting log-normal models accounting for interval censoring.

**Results:** We identified 115 relevant papers and obtained data from 66 (57.4%) – representing a total of 1198 patients across 14 countries. SARS-CoV-2 viral load peaks prior to symptom onset and remains elevated for up to three weeks, while MERS-CoV and SARS-CoV viral loads peak after symptom onset. SARS-CoV-2, MERS-CoV, and SARS-CoV had median viral shedding durations of 4.8, 4.2, and 1.2 days after symptom onset. Disease severity, age, and specimen type all have an effect on viral load, but sex does not.

**Discussion:** Using a pooled analysis of the largest collection of IPD on viral load to date, we are the first to report that SARS-CoV-2 viral load peaks prior to – not at – symptom onset. Detailed estimation of the trajectories of viral load and virus shedding can inform the transmission, mathematical modeling, and clinical implications of SARS-CoV-2, MERS-CoV, and SARS-CoV infection.

## Introduction

The emergence and pandemic spread of Severe Acute Respiratory Syndrome coronavirus 2 (SARS-CoV-2) in 2019-2020 has warranted rapid research to understand the risk factors driving transmission and severe morbidity. To date, 216 countries have been affected worldwide,[1] with considerable geographic heterogeneity in terms of reported case counts and case fatality.[2] Policies, detection capacity, and behavioral decisions have contributed to heterogeneity at national and regional scales.[3, 4, 5] The effectiveness of mitigation efforts are inextricably linked to the viral dynamics that underlie infection progression and transmission potential and that delineate subpopulations of greatest epidemiological significance (e.g., asymptomatic and presymptomatic patients).[6, 7]

Across pathogens, viral load can serve as an indicator of clinical outcome [8, 9, 10, 11] and a metric of transmissibility.[12, 13] For coronaviruses specifically, individual studies have suggested that viral load and infectiousness peak during the presymptomatic phase for COVID-19[14] and after symptom onset for SARS and MERS,[15, 16] and that higher viral loads may signal heightened risk of mortality,[7, 9, 15, 17] after accounting for age and other correlates. Such data provides key parameters that inform and enhance the appropriate structure and predictive power of mathematical models.[18]

Mathematical modeling has importantly served to quantify potential outbreak size for emerging coronaviruses both in the absence of mitigation and after introduction of intervention.[19, 20, 21] Models that include subpopulation-specific viral dynamics can more accurately assess the relative contributions to transmission of individuals in asymptomatic, presymptomatic, and symptomatic states and across age, sex, and comorbid status. During the COVID-19 crisis, this information on transmission is important in order for models to be used to guide recommendations around social distancing policies, including school closures, quarantine and isolation, and to project potential healthcare capacity needs.

Aggregation of data across studies allows for improved power to statistically describe viral dynamics overall according to epidemiologically relevant subgroups. To date, researchers have systematically reviewed published studies on viral dynamics for coronaviruses (MERS-CoV, SARS-CoV-2) and provided metrics on pooled parameters, such as duration of viral shedding, across different sampling sites.[7, 22, 23, 24] However, aggregation of individual-level data across studies has not been undertaken on this scale. Here we provide pooled time-series distributions of coronavirus viral load and the duration of viral shedding across patient subgroups defined by demographic and clinical characteristics, after controlling for study-specific methodologies. We define viral load as quantitative viral titre (e.g., copy number) and viral shedding as the qualitative description of disease status (e.g., PCR positve). The distributions and associated parameters are intended to better understand the transmission dynamics of SARS-CoV-2 and inform mathematical models for improved investigation of population-specific mitigation strategies.

## Methods

### Search Strategy

On June 8th, 2020, the investigators performed a PubMed search with the phrase, “((corona*) OR (COVID*) OR (”SARS-CoV-2”) OR (SARS*) OR (MERS*)) AND ((”viral load”) OR (”viral shedding”) OR (serolog*)).” We used PubMed’s online advanced search algorithm. We did not specify a category or tag (e.g., “Year” or “Author”) for any of these terms, so the Boolean phrase was searched across all fields. We did not limit our search results by publication date.

### Study Selection

Each title was blindly screened by three reviewers. Titles were only removed if all three reviewers agreed that they were not broadly pertinent to the study objective of assessing viral dynamics of coronaviruses in humans (Figure 1). If at least one reviewer selected a title, then it was included in abstract screening. Each abstract was read by at least two reviewers and was categorized as one of the following: (1) study of temporal dynamics of viral load of human coronavirus; (2) study of human coronavirus but not of temporal dynamics of viral load; (3) study in non-humans or basic biological study of viral load that did not involve humans; (4) study in humans not about coronavirus; (5) not in English; (6) duplicate of another record; (7) duplicate data from another paper; or (8) dead link/bad return/noise. If there was disagreement about the categorization, a third reviewer acted as a tie-breaker.

**Figure 1:**
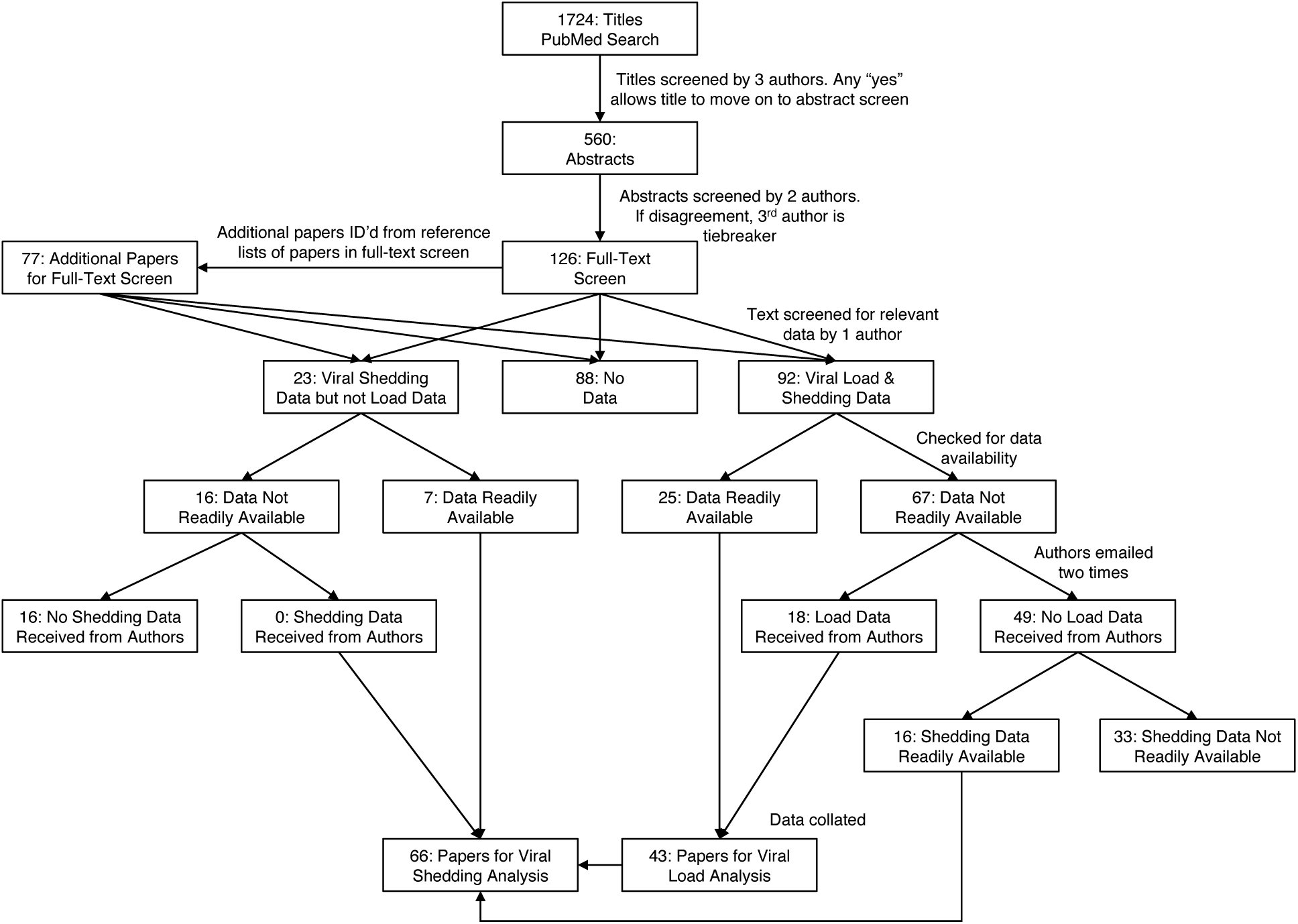
Summary of systematic review. A search of PubMed sources identified 1724 records. After title and abstract screening, 203 titles underwent full-text screen. Forty three and 66 studies met the inclusion criteria and were included in the systematic review for viral load and shedding analysis, respectively.

Abstracts that were categorized as a study of temporal dynamics of viral load of human coronavirus were then included in a full-text screening by at least one reviewer. The reference lists of these papers were manually scanned for additional relevant studies that were automatically added to the list of papers requiring a full-text screen. During the full-text screen, we included papers that (1) had novel viral load data or (2) had novel data on the duration of viral shedding. We included papers that mentioned these types of data even if the data was not directly presented in any way. We did not exclude drug trials or case studies as long as the studies contained at least one data point (i.e., including both Time and either Viral load or Shedding). We included studies with any temporal measurement (time since: symptom onset, hospital admission, diagnosis, treatment, etc.) and any specimen type (e.g., nasopharyngeal, sputum, stool), and we included pre-prints and letters to editors. We excluded papers that (1) reported no novel viral load or viral shedding data or (2) estimated viral load with serum titre, because it has been shown that seroconversion does not occur until around 10 days after symptom onset.[25]

When individual-level data were not openly available (printed (non-estimated) number somewhere in the text, a table, a figure, or a supplement) with the published articles selected via full-text review, we contacted corresponding authors through email to request data access. We made up to two attempts to connect with authors. Papers marked as having novel viral load data for which we did not receive access were screened once more for readily available viral shedding data. We took several measures to reduce the risk of bias in our methods and in our included studies (details in Supplementary Appendix A).[26, 27]

### Data Extraction

Data extraction was performed by three authors (AEB, AC, and RAA), and three data points from each paper were randomly checked for data entry or interpretation errors by one author (LAS). For each paper or associated dataset, we extracted viral load data (copies or cycle threshold values [Ct] value) and/or viral shedding data (positive or negative) relative to time (days) at the patient level. Further, we extracted study-specific data, assay-specific data, and additional patient-specific data. The following study-specific fields were extracted: (1) title, (2) disease, (3) the start and end dates of the study, (4) the location (city/region and country), and (5) the reference point of temporal measurement (symptom, admission, etc.). Each study was assigned a Paper ID. If the paper had time measured with respect to multiple reference points (e.g., since symptom onset and since admission), both were extracted. Days were scaled so that the first day of symptoms was equal to day zero. Where applicable, the following assay-specific data were extracted: (1) the Ct cutoff value for positive and negative results, (2) the type of PCR being used, (3) the PCR target gene sequence, and (4) the specimen type. All specimen types were classified into four categories: respiratory, gastrointestinal, urinary, or serum. Finally, anonymized patient IDs were extracted, as were the following patient data (when available): (1) age, (2) sex, (3) disease severity (mild = symptomatic, moderate = hospitalized, severe = requiring ventilation, intubation or death),(4) hospitalization status, and (5) whether the patient received treatment.

### Statistical analysis

The temporal measurements were scaled to days since symptom onset using existing data (see Supplementary Appendix B). For example, if the paper measured time in days since exposure, then we subtracted the median incubation period from the temporal measurement. Thus, 20 days since exposure was scaled to 12 days since symptom onset using the current median incubation period, 7.7 days, rounded to the nearest integer.[28]

We transformed viral load data that was reported in Ct values to log_10_ Copies / ml using an average standard curve (see Supplementary Appendix C). The variation that was introduced by these two scaling methods was mitigated by treating the Paper ID as a random effect in our models. We assumed that negative tests between multiple positive tests were false negatives. We only kept negative test results that were the last negative test before the first positive test or the first negative test after the last positive test (i.e., we removed leading or trailing zeros).

Bivariate comparisons were conducted to evaluate associations between demographic (i.e., age and sex) and clinical characteristics (i.e., hospitalization status) of patients, and between these characteristics and viral load measurements, regardless of time since symptom onset. Analysis of variance (ANOVA) was conducted to assess whether the distribution of viral load measurements significantly differed over specimen type, disease severity, patient sex, and patient age categories, by disease.

A series of generalized additive models (GAM) were used to explore different aspects of viral load kinetics. First, to assess SARS-CoV-2 viral load dynamics, we used a GAM with viral load (log_10_ copies/ml) as the dependent variable, symptom days as the main predictor variable, paper ID as a random effect, and patient age, patient sex, disease severity, specimen category, and PCR gene target as covariates (Model 1, Table 2). To address missing data in individual variables, we imputed 29.8% of age data, 29.7% of sex data, 36.6% of severity data, and 12.1% of PCR gene target data using multivariate imputation by chained equations (R MICE package, v 3.11.0, sensitivity analyses of imputations can be found in the supplementary materials, Appendix D).[29] When imputation was used, we performed five parallel imputations with 20 iterations each. Model estimates and predictions were from the pooled parallel imputations. Second, to investigate gastrointestinal and respiratory viral load trajectories, we adapted the above model to only include those two specimen categories which we treated as the main factor in the model (Model 2). Finally, to quantify differences in respiratory viral load kinetics across diseases (SARS, MERS, and COVID-19), we simplified our model, because SARS and MERS had fewer complete datasets (e.g., missing age or sex fields). We included only respiratory specimens, and our GAM treated symptom days as the main predictor variable, disease as the main factor, PCR gene target as a covariate, and paper ID as a random effect (Model 3). Here, we imputed 11.8% of gene data (sensitivity analyses in supplementary materials, Appendix E).

### Duration of shedding

A survival analytic approach was used to determine duration of viral shedding. Data were classified as fully observed, single- or doubly-interval censored. Observations were fully observed if tested samples had a negative test before and after the first and last positive test, respectively. If samples were found to be positive on the first or last sample taken, then the data point was assumed single-interval censored. If samples were positive on both the first and last sample, then the data point was assumed doubly-interval censored. Missing observations or a negative surrounded by two positive samples were assumed to be positive. Methods for analyzing doubly-interval censored data have been developed previously [30, 31]. We verify the duration of virus shedding to be log-normally distributed (see Supplementary Material, Appendix F).

## Results

Our PubMed search yielded 1,724 results (Figure 1). Following title-screening, we read 560 abstracts, and we then performed a full-text screen on 203 papers. The full text screen resulted in 92 papers with viral load data, 23 with viral shedding but not viral load data, and 88 with no relevant data. We emailed authors regarding 89 papers, and received data from 18. There were 16 papers for which we did not receive access to viral load data that did have viral shedding data readily available. If patients had a positive viral load, then they were also positive for viral shedding, so all viral load papers were included as viral shedding papers. Ultimately, we included 43 papers in our analysis of viral load distribution and 66 in our study of the duration of viral shedding (see supplementary Appendix G for table of papers used).[25, 32, 33, 34, 35, 36, 37, 38, 24, 39, 40, 41, 42, 43, 44, 45, 46, 47, 48, 49, 50, 51, 52, 53, 54, 55, 56, 57, 58, 59, 60, 61, 62, 63, 64, 65, 66, 67, 68, 69, 70, 71, 72, 73, 74, 75, 76, 77, 78, 79, 80, 81, 82, 83, 84, 85, 86, 87, 88, 89, 90, 91, 92, 93, 94, 95] The included papers yielded 932 unique individuals with 5328 observations for the viral load analysis and 1198 individuals and 7240 observations for the viral shedding analysis (Table 1). SARS-CoV-2 was the most common virus for which we had data, accounting for 77% of patients in both the viral load and the viral shedding datasets. China, USA, Taiwan, and South Korea were the most common locations of patients included in the dataset. As for individual-level characteristics, median ages were 44 and 43 for viral load and shedding, respectively and 62% female for both. Finally, the majority of cases in each analysis group were of mild severity and not hospitalized. Age, sex, severity, and hospital statistics are based primarily on COVID-19 cases, for which there was the most complete data.

**Table 1:** Paper and Patient Statistics. Table shows summary statistics for papers, patients, and tests used in our analyses. *‡* and © denote Chi-squared and t-tests for differences between groups, respectively. Two patients had different specimens measured in both log_10_ Copies and in CT values (Both). Everything but Papers and Observations represent patient counts. Because we did not have complete data for age, sex, severity, and hospitalization status, the number of individuals will not sum to 932 for viral load or 1198 for shedding.

|  | Viral load | Shedding | Test |
| --- | --- | --- | --- |
| Papers | 43 | 66 |  |
| Individuals | 932 | 1198 |  |
| Observations | 5328 | 7240 |  |
| SARS-CoV-2 | 715 (76.7%) | 917 (76.5%) | p = 0.199‡ |
| MERS | 38 (4.1%) | 72 (6%) |  |
| SARS | 179 (19.2%) | 209 (17.4%) |  |
| China | 376 (40.3%) | 537 (44.8%) | p = 0.22‡ |
| USA | 291 (31.2%) | 353 (29.5%) |  |
| Taiwan | 154 (16.5%) | 155 (12.9%) |  |
| South Korea | 47 (5%) | 52 (4.3%) |  |
| Singapore | 41 (4.4%) | 41 (3.4%) |  |
| Others | 23 (2.5%) | 60 (5%) | p = 0.471§ |
| Age | 44.0 (IQR: 31.0–59.0) | 43.0 (IQR: 30.0–59.0) |  |
| Female | 299 (61.9%) | 337 (61.5%) | p = 0.74§ |
| Male | 184 (38.1%) | 211 (38.5%) |  |
| Mild | 266 (64.9%) | 269 (50.1%) | p = 0.199‡ |
| Moderate | 102 (24.9%) | 192 (35.8%) |  |
| Severe | 42 (10.2%) | 76 (14.2%) |  |
| Not Hospitalized | 266 (64.9%) | 269 (50.1%) | p = 0.47§ |
| Hospitalized | 144 (35.1%) | 268 (49.9%) |  |
| Both | 2 (0.2%) |  |  |
| CT | 290 (31.1%) |  |  |
| ΔCT | 31 (3.3%) | | |
| log<sub>10</sub> Copies | 609 (65.3%) |  |  |

Our bivariate analyses showed that viral loads were significantly negatively associated with age and disease severity, and that females had significantly higher viral loads than males. These two patterns did not exist when controlling for the time of the test, relative to symptom onset. Increasing disease severity was significantly associated with age and sex. There is an average age difference of 22.0 years between mild and severe cases, 19.5 years between severe and moderate cases, and 2.5 years between mild and moderate cases (Tukey test p-values = 0, 0.00166, and 0.3422, respectively). On average, patients who were hospitalized are 5.1 years older than those who were not hospitalized (p-value=0.0096). Males have more severe disease than females (p-value= 1.364 e-07), and they are more likely to be hospitalized (p-value = 1.504e-07).

### Viral Load

Model 1 shows that SARS-CoV-2 viral load peaks prior to symptom onset and decreases over time with the steepest decrease occurring between days 1 and 11 after symptom onset (p-value = 2e-16, Table 2, Figures 2, 3). On average, the smoothed splines for age trended negatively. There was a strong negative association between increasing age and viral load for ages *≤* 16 and for ages *≥* 85, but viral loads between ages 16 and 85 were relatively constant (p-value = 8.3e-7). There were no sex differences in viral load, nor were there major differences between mild and moderate patients. However, severe patients had significantly higher viral loads than mild patients (9.9% higher after converting from log_10_ copies to raw copy number). All specimen categories had a significant effect on viral load, with gastrointestinal specimens having the highest loads and urinary specimens having the lowest (p-values = 7.3e-05 and 5.8e-03, respectively, Table 2).

**Table 2:**
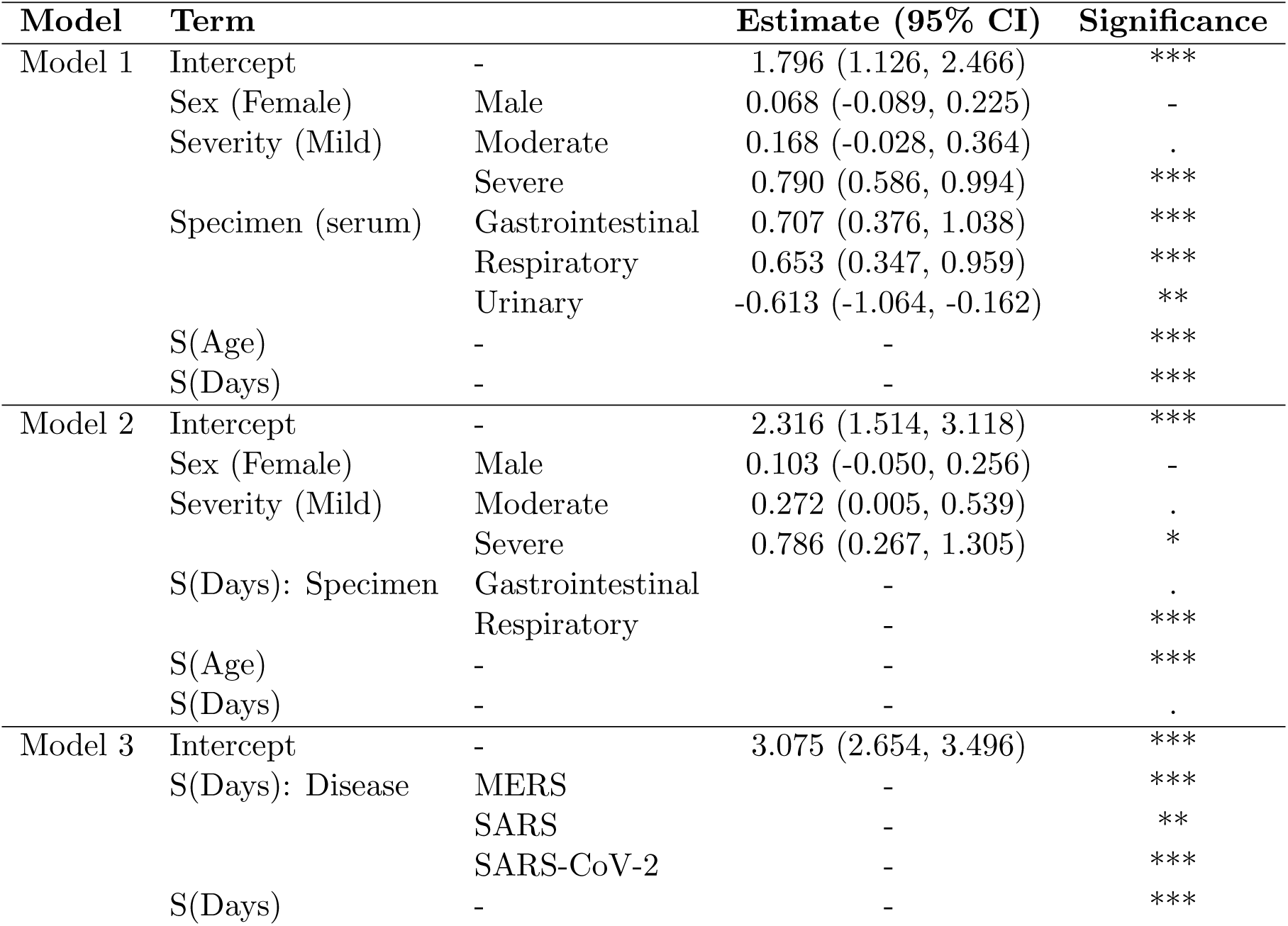
Models. Table shows model estimates, 95% confidence intervals, and the significance of those terms for the three models we used to assess viral load dynamics in coronaviruses (sig. codes: 0 = ***, 0.001 = **, 0.01 = *, 0.05 = ., >0.05 = -). Model 1 and 2 represent only SARS-2 data, and Model 3 compares across diseases. Terms without an estimate are smoothed terms (over days or age). Therefore, they have multiple coefficients each corresponding to an individual spline, which is why no single value can be presented here.

**Figure 2:**
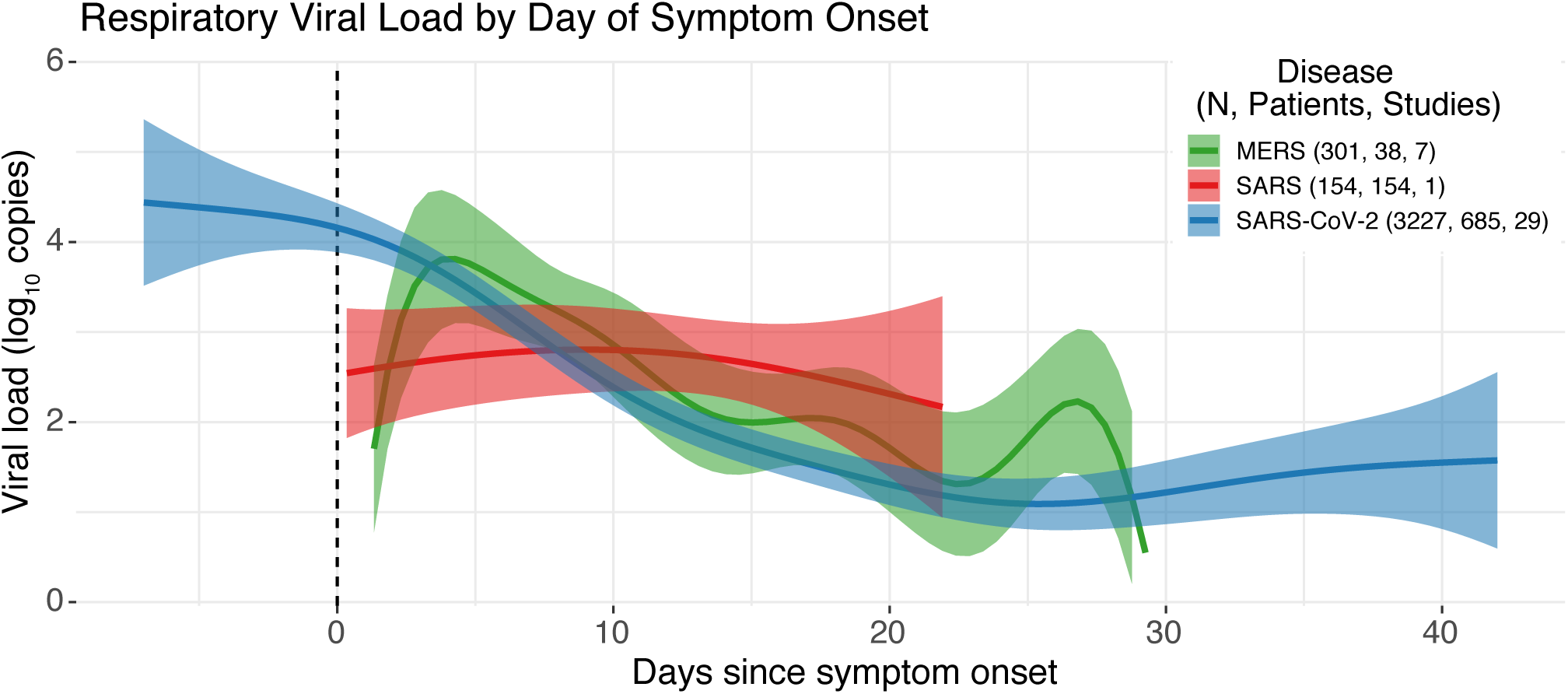
SARS-CoV-2, MERS-CoV, and SARS-CoV viral loads over time. Figure shows estimates of the three pathogen viral loads over time from the adjusted GAM model.

**Figure 3:**
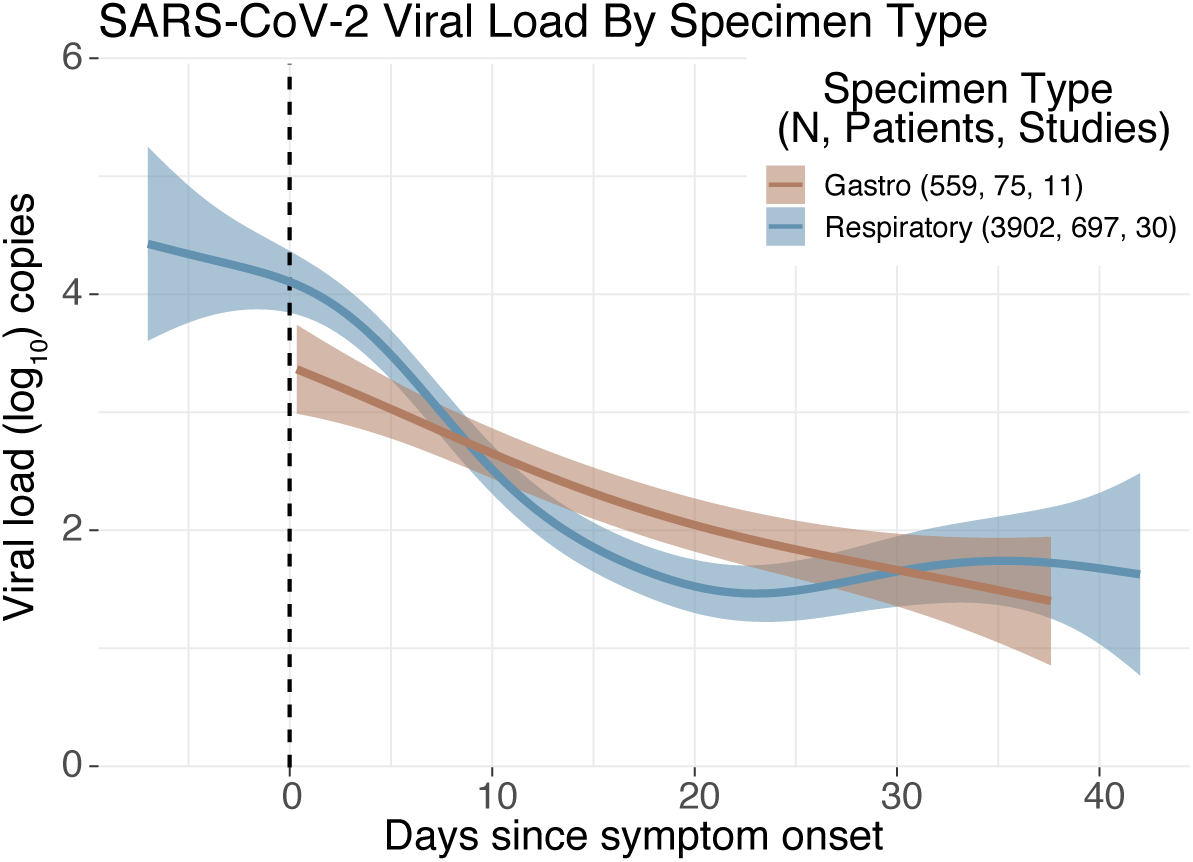
Comparison of SARS-CoV-2 viral loads between respiratory and gastrointestinal sampling. Figure shows estimates of SARS-CoV-2 viral loads over time from the adjusted GAM model comparing respiratory and gastrointestinal sampling.

Model 2 explores differences in SARS-CoV-2 gastrointestinal and respiratory viral load over time. This model treats the two smoothed specimen categories (only) as main factors (Figure 3). In this model, the smoothed effect for days was only weakly significant (p-value = 0.06). The smoothed effect for age trended strongly negative for ages *≤* 16, but in this model ages *≥* 75 were positively associated with increasing viral load (p-value = 2e-16). Severe patients had significantly higher viral loads than mild patients (3% higher, p-value= 2.79e-02). The smoothed effect of gastrointestinal specimen type over days was not significant. It had 1.03 effective degrees of freedom (EDF), indicating that it has a linear relationship with days and was therefore insignificant when smoothed over days. In contrast, respiratory specimens were very significant (5 EDF, p-value = 2e-16). Viral load in respiratory specimens decreased constantly from 7 days prior to symptom onset to 23 days afterward, with the sharpest decline occurring between days 1 and 22 after onset. Between days 22 and 35, we observed a slight increase in viral load, which then plateaus around day 35. Sex and mild versus moderate severity were insignificant and weakly significant, respectively (p-values = 0.219 and 0.058).

Finally, Model 3 compares the respiratory viral load kinetics of SARS-CoV, MERS-CoV, and SARS-CoV-2 by treating disease as the main factor, smoothed over days (Figure 2). All model estimates were significant. MERS smoothed over days had 8.7 EDF (p-value = 6.54e-06). For MERS, viral load increases until day 3 after symptom onset, and then decreases consistently. SARS-1 peaks at or shortly after symptom onset and then decreases nearly linearly (EDF = 1.38, p-value = 0.0036). Finally, SARS-CoV-2 peaks prior to symptom onset, and then decreases until around day 25 when there is a slight uptick (EDF=5.5, p-value = 2e-16).

### Duration of shedding

Durations of viral shedding in respiratory samples were similar for SARS-CoV-2 and MERS-CoV with median durations of 4.76 days (95% CI: 3.4.4 to 5.11) and 4.12 days (95% CI: 3.25 to 4.99), respectively (Figure 4 and Table 3). These durations were significantly longer than for SARS-CoV which had a median duration of 1.23 days (95% CI: 1.1 to 1.36). In gastrointestinal samples, SARS-CoV-2 had significantly longer durations of shedding than MERS-CoV, with medians of 4.94 days (95% CI: 4.09 to 5.8) and 1.85 days (95% CI: 0.6 to 3.1), respectively.

**Table 3:** Estimated durations of viral shedding for SARS-CoV-2, MERS-CoV, and SARS-CoV. Table shows the days of shedding for the three viruses in respiratory and gastrointestinal samples as estimated using a time to even survival analysis approach to adjust for censored data (see [30, 31]). N indicated the number of subjects included in each category.

| Respiratory shedding |  |  | Gastrointestinal shedding |  |  |
| --- | --- | --- | --- | --- | --- |
| Virus | Percentile | Days (95% CI) | Virus | Percentile | Days (95% CI) |
| SARS-CoV-2<br>n = 1300 | 5th | 0.52 (0.46, 0.58) | SARS-CoV-2<br>n = 198 | 5th | 0.71 (0.53, 0.90) |
|  | 25th | 1.92 (1.76, 2.07) |  | 25th | 2.24 (1.82, 2.65) |
|  | 50th | 4.76 (4.40, 5.11) |  | 50th | 4.94 (4.09, 5.80) |
|  | 75th | 11.81 (10.82, 12.81) |  | 75th | 10.93 (8.79, 13.07) |
|  | 95th | 43.73 (38.62, 48.85) |  | 95th | 34.22 (24.90, 43.54) |
| MERS<br>n = 135 | 5th | 0.55 (0.37, 0.72) | MERS<br>n = 7 | 5th | 0.56 (-0.02, 1.13) |
|  | 25th | 1.80 (1.38, 2.22) |  | 25th | 1.13 (0.28, 1.97) |
|  | 50th | 4.12 (3.25, 4.99) |  | 50th | 1.85 (0.60, 3.10) |
|  | 75th | 9.43 (7.21, 11.65) |  | 75th | 3.02 (0.76, 5.29) |
|  | 95th | 30.98 (20.86, 41.11) |  | 95th | 6.15 (-0.23, 12.53) |
| SARS<br>n = 158 | 5th | 0.40 (0.34, 0.47) | SARS<br>n = 0 | — |  |
|  | 25th | 0.78 (0.68, 0.87) |  |  |  |
|  | 50th | 1.23 (1.10, 1.36) |  |  |  |
|  | 75th | 1.95 (1.72, 2.18) |  |  |  |
|  | 95th | 3.78 (3.16, 4.41) |  |  |  |

**Figure 4:**
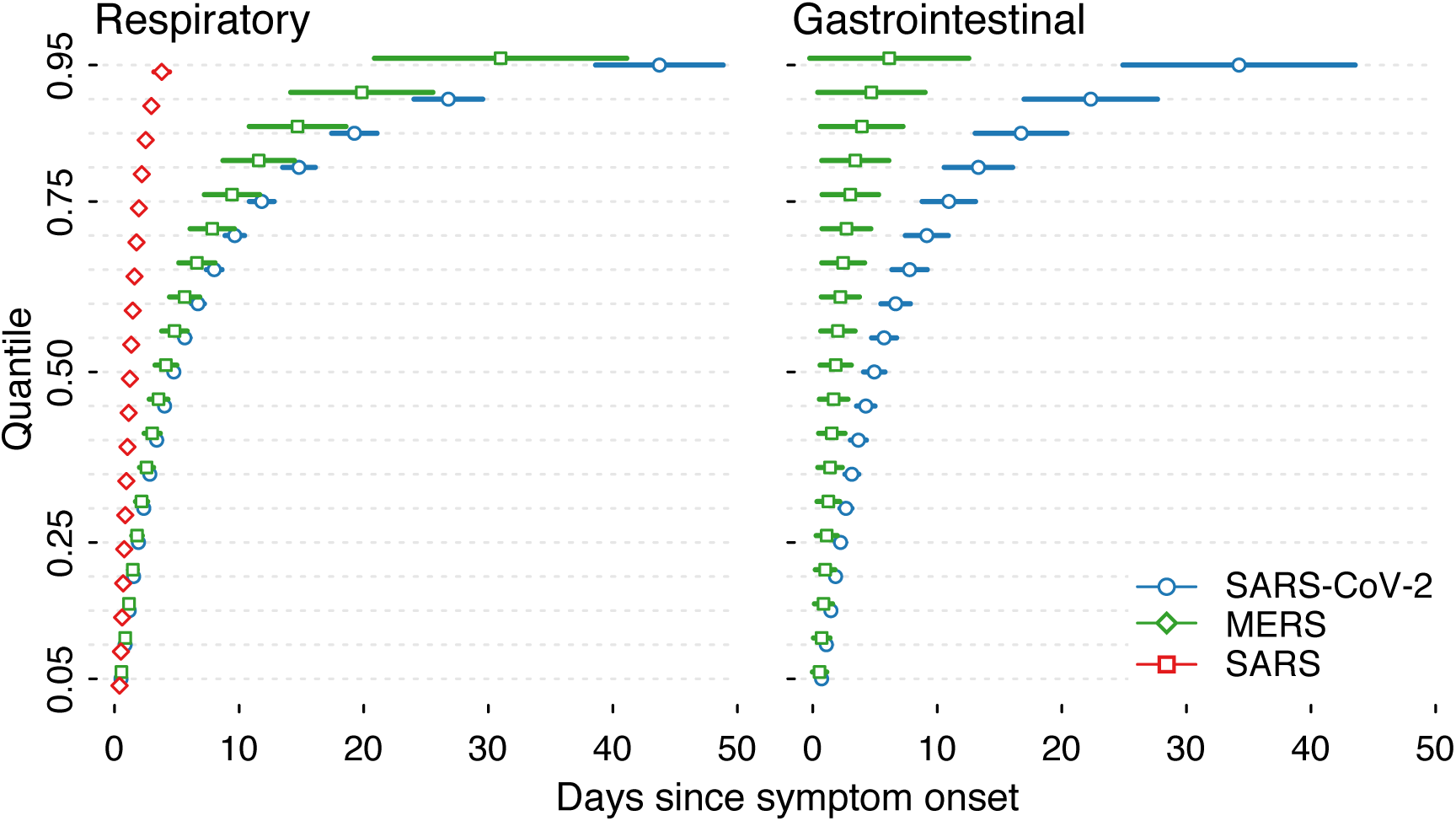
SARS-CoV-2, MERS, and SARS viral shedding over time. Figure shows the log-normal estimates of viral shedding for the three diseases with both gastrointestinal and respiratory samples.

## Discussion

As SARS-CoV-2 circles the globe killing thousands, sickening millions, and stretching hospital capacities, mathematical modeling of the transmission dynamics has proven invaluable for the prediction of new cases of COVID-19 for decision-making by medical and public health professionals and policy-makers [96, 97, 98, 99, 100]. Recent work has explored the duration of SARS-CoV-2 viral shedding [101, 102, 23], and the role of asymptomatic and presymptomatic shedding [103], but as of yet, no systematic review has pooled individual-level data to quantitatively describe viral load dynamics of the three emergent human coronaviruses across the disease course. Here we present temporal viral load trajectories for 932 symptomatic cases from 43 studies, and viral shedding data for 1198 patients from 66 studies, conducted in 14 countries. We find that viral load dynamics differ across coronaviruses; while SARS-CoV-2 peaks before symptom onset and continues to remain elevated for up to three weeks after onset, SARS-CoV and MERS-CoV peak after symptom onset. SARS viral load is lower at all daily time steps after symptom onset, as compared to viral load for MERS and SARS-CoV-2. While our results are consistent with individual studies and other reviews [48, 22, 25], our use of individual-level data allowed for statistical modeling to quantify temporal distributions with adjustment for clinical and demographic covariates.

We examined differences in viral load and shedding across a range of covariates. No differences in duration of SARS-CoV-2 viral shedding were seen by sex or with age, although the smoothed effect for age was statistically significant with a negative association, however further study is warranted. There were statistically significant differences comparing severe cases to mild, with mild cases having only 16% of the viral load of severe cases. While there may be some selection bias with those with severe disease being hospitalized and possibly tested more frequently, we included random effects to account for variability between studies and sampling pools.

Recent studies have shown the presence of SARS-CoV-2 in fecal samples [92, 104, 105] such that environmental surveillance using wastewater has been recommended [106, 107, 108, 109]. We find comparable viral load trajectories between respiratory and gastrointestinal samples, with a more gradual decline in load in gastrointestinal samples, and nearly identical durations of shedding. This implies that using wastewater for surveillance is likely to be a valid tool for detecting cases for the full duration of illness, and subsequently, transmission. While, it should be noted that some have argued that viral RNA in feces might not represent live virus [24], more research is needed to confirm this.

Several mathematical models of SARS-CoV-2, MERS, and SARS have incorporated viral load dynamics into formulations of their models [110, 111]. As has been presented here, the individual-level viral dynamics of SARS-CoV-2 show a clear pattern of a peak in viral load at or before the appearance of symptoms and a steady log-fold decline over the next 21 days. This is distinct from MERS where peak viral load occurs several days after symptom onset; ignoring these important differences would make predictions from simpler models incorrect with potentially dangerous public health responses. Summarized data and raw data from the identified studies presented here could prove useful to parameterizing more complex models with realistic viral dynamics. In particular, curves predicted using the GAM models offer temporal distributions that could be incorporated into models to reflect changes in infectiousness over the course of infection, with or without consideration for sex, age, and severity status depending on the model structure.

It has been suggested that PCR positivity does not correspond to transmission potential as successful culture has not been achieved beyond 8 days after the onset of symptoms and because PCR positivity has been observed for up to 60 days.[25, 112, 72, 24] While PCR positivity can arise from either live virus or viral fragments (particularly in feces), viral culture is notoriously difficult, and thus, we disagree with the idea that the absence of successful culture implies the absence of live virus or transmission risk.[48]

The ongoing pandemic has seen an unprecedented [113] display of open science [114, 115]. There has been widespread sharing of genetic sequences, code, data, and article preprints. This commendable – and hopefully permanent – advance in medicine and epidemiology has facilitated more rapid and cross-disciplinary science to rapidly improve understanding around how to mitigate morbidity and mortality due to SARS-CoV-2. In conducting our systematic review we found that only 59% and 27% of papers had freely available viral shedding and load data (either published with the study or on an open data archive). Of the 89 authors emailed to request data access, 43% responded and only 20% responded with data. More stringent standards for data availability statements would provide authors with opportunities to explicitly clarify what is and is not available to share and elucidate why, such as concerns around patient confidentiality. To keep the strong culture of open science moving forward, we, as scientists, need to ensure that data sharing is a norm, not an exception. As other groups are undertaking’living’ systematic reviews and meta-analyses with study-level data, it is our goal to incorporate ongoing, active contributions into our pooled dataset. This data will be made available for clinicians, public health decision-makers, and modelers to use for data-driven insights on transmission.

This, like every study, is not without limitations. First, 65% of the studies included reported only Ct values from the qPCR samples. Copy number (32% of our data) is a standardized measure of viral load across studies whereas Ct values are not. To compare across papers, we had to convert Ct values to log_10_ copy number using an average standard curve. This introduced artificial variation in our data, which we controlled for by treating study ID as a random effect. We also ran a sensitivity analysis comparing the published, standardized copy numbers to our copy-number converted Ct values (Supplementary Appendix C). Though we observed no major qualitative differences between the raw and converted data (especially within diseases), the value of the peak viral loads (in log_10_ copies) that we report here should not be taken out of context. Having used standard methods of conversion, this leads to a limitation of the field writ large. Second, limited numbers of MERS fecal shedding and none for SARS limits the ability to draw firm conclusions about the transmission routes of these pathogens. However these estimates do suggest lines of enquiry for future research. Third, due to incomplete recording of data, we had to impute missing values to increase statistical power. This did not qualitatively change the viral load trajectories (Supplementary Appendix D and E) and is common practice when dealing with diverse observational data. Fourth, due to the novelty of each coronavirus when it emerged, many of the original studies were case studies with patients not being representative of the population at large. However, by definition, case studies will have little statistical influence over larger cohort studies. Finally, due to a lack of uniformity in how days were numbered we had to make assumptions for the timing of symptom onset. Sensitivity analyses have shown that these assumptions do not make qualitative differences in the results, and we applied a consistent timing scheme within the dataset.

The current study is to the best of our knowledge the only study that has pooled individual-level temporal viral load dynamics and shedding duration across studies for all three emergent human coronaviruses, including the novel coronavirus, which is responsible for the ongoing pandemic. Our analysis provides robust evidence in support of findings to date that viral load for SARS-CoV-2 peaks during the presymptomatic phase. This has strong policy implications for the surveillance and control of SARS-CoV-2, and for the ongoing reopening of businesses, schools, and borders globally. The implications of transmission during the presymptomatic phase – possibly, the period of highest transmissibility given the viral load dynamics presented here – indicate the need for strong contact tracing, both forward and backward [116], isolation measures, and in turn, buy-in of individuals and communities in such efforts. Without appropriate intervention, presymptomatic and asymptomatic transmission could lead to ongoing waves of resurgence, as has been suggested to explain the current resurgence of whooping cough in the US and elsewhere [117]. Through the disruption of situational awareness and potentially substantial undocumented transmission, hospitals, schools, and businesses face difficult decisions about staffing and opening in general. Careful consideration towards premature opening of schools and non-essential businesses demands attention in the face of this looming threat. [118]

## Data Availability

Data will be made available after publication of the manuscript.

## Acknowledgements

We are particularly grateful for the assistance given by Aaron Westmoreland in debugging code and for his consistent feedback, and for helpful comments from Edward Wenger. We are appreciative of authors of papers that had data readily available. Further, we’d like to thank Drs. Leo Poon, Kelvin To, Y.M. Dennis Lo, Quang D. Pham, Nabin K. Shrestha, Qingfeng Sun, Clemens Wendtner, Victor Corman, Yuan Xue, Wai K. Leung, Nam-Hyuk Cho, Myoung-don Oh, John Jernigan, Quan-Sheng Xing, Gary Wing-Kin Wong, Yuhan Xing, Bin Wu, Kai-qian Kam, Mei-Shang Ho, Mona Loutfy, Claire M. Midgley, and the US CDC COVID-19 Investigation Team for kindly providing data. AEB acknowledges support from an internship at the Institute for Disease Modeling. This publication is based on research developed by the Institute for Disease Modeling prior to its affiliation with the Bill Melinda Gates Foundation. The findings and conclusions contained within are those of the authors and do not necessarily reflect the official positions or policies of the Bill Melinda Gates Foundation. For BMA, LAS, and SC, this work was supported by the Global Good Fund.

